# Decomposing the Caesarean section trend in Bangladesh (2012–2025): A multivariate Linear Probability Model analysis

**DOI:** 10.64898/2026.10.28.26366781

**Authors:** Md Akhtaruzzaman Limon

## Abstract

**Background:** While Caesarean section is a life-saving intervention when medically indicated, the World Health Organization recommends an optimal population-level rate of 10% to 15% to prevent iatrogenic morbidities. In Bangladesh, however, rapid clinical integration within a commercialized, dual-practice healthcare architecture has coincided with an unprecedented escalation in surgical deliveries. Because standard cross-sectional models cannot algebraically separate population shifts from changing provider propensity, this study aimed to decompose the compositional and behavioral drivers of the Caesarean section surge in Bangladesh between 2012 and 2025.

**Methods:** Nationally representative data were analyzed from the 2012–2013, 2019, and 2024–2025 Bangladesh Multiple Indicator Cluster Surveys (MICS), comprising 26,534 women aged 15–49 years who delivered a child within the preceding two years. Survey skip-logic was harmonized to retain unmedicalized cohorts receiving zero antenatal care (ANC). A survey-weighted multivariate Linear Probability Model (LPM) Oaxaca-Blinder decomposition was applied to a complete-case cohort (N=16,179) to partition the prevalence change into population-level compositional shifts (endowments) and behavioral changes (coefficients).

**Results:** National Caesarean section prevalence surged by 32.47 percentage points, rising from 19.48% in 2012 to 36.05% in 2019 and 51.95% in 2025 (p<0.001). Surgical delivery multiplied over sixfold among the poorest wealth quintile (5.53% to 34.37%), rose by 34.83 percentage points among rural mothers (15.66% to 50.49%), and reached 85.12% within private facilities. Multivariate decomposition demonstrated that 87.77% of the overall increase was attributable to compositional shifts rather than behavioral factors (12.23%). The sharp reduction in pregnancies receiving zero ANC visits (35.52% to 8.12%) independently drove 49.04% of the surge, while increased utilization of private clinics and NGOs (18.20% to 54.36%) explained 43.39%. Conversely, biological indications such as fetal macrosomia contributed less than 1%.

**Conclusions:** The unprecedented escalation of Caesarean sections in Bangladesh represents a profound pattern of systemic over-medicalization that is independent of baseline clinical risk. The surge is overwhelmingly structural, driven by the disproportionate concentration of deliveries within the commercialized private healthcare sector and supplier-induced demand following integration into formal clinical pathways. Reversing this trajectory necessitates immediate regulatory intervention, specifically the mandate of clinical auditing frameworks such as the Robson classification, to decouple financial incentives from obstetric decision-making in private facilities.

## Introduction

It has long been recognized that Caesarean section is a vital, life-saving surgical intervention necessary to reduce maternal and neonatal mortality in the presence of strict obstetric indications, such as prolonged labor, fetal distress, or abnormal presentation [1,2]. To optimize population-level maternal and perinatal outcomes, the World Health Organization (WHO) recommends that national Caesarean section rates should optimally reside between 10% and 15% [1]. However, when performed without medical justification, surgical deliveries expose mothers and infants to significant iatrogenic risks, including postpartum hemorrhage, systemic infection, extended recovery periods, and an elevated probability of uterine rupture or abnormal placentation in subsequent pregnancies [3,4]. Beyond direct clinical morbidities, the routine overuse of Caesarean sections imposes a substantial financial burden on healthcare systems and exacerbates catastrophic out-of-pocket health expenditures, particularly in resource-constrained low-and middle-income countries (LMICs) [5].

Globally, the prevalence of Caesarean deliveries has escalated steadily over the past two decades [6]. This trend is acutely pronounced in LMICs undergoing the “obstetric transition”, which is a systemic shift characterized by the rapid migration of deliveries from home environments to institutional clinical settings [7]. In Bangladesh, decades of targeted public health interventions and demographic shifts have dramatically expanded maternal healthcare coverage. Yet this integration into formal clinical pathways has been accompanied by a steep, paradoxical increase in surgical deliveries. Historical analyses utilizing demographic surveys documented an increase in the national Caesarean rate from approximately 4% in 2004 to over 33% by 2017 [8,9].

Understanding the drivers of this escalation requires examining the localized healthcare architecture. In Bangladesh, the expansion of facility-based deliveries has been heavily absorbed by the private healthcare sector, operating within a highly commercialized, fee-for-service environment characterized by fragmented regulatory oversight [10]. This dual-practice healthcare system structurally facilitates supplier-induced demand, where financial and operational incentives disproportionately favor surgical interventions over vaginal births [11]. Previous determinants analyses indicate that Caesarean deliveries in Bangladesh were historically concentrated among urban, highly educated, and affluent populations utilizing these private facilities, reflecting severe information asymmetry between providers and patients [12].

While existing cross-sectional literature successfully identifies individual-level predictors of surgical delivery, significant methodological and temporal gaps remain. Standard epidemiological approaches predominantly utilize logistic regression models at single time points to generate odds ratios for discrete risk factors [12]. These models are mathematically constrained from isolating the structural determinants of temporal change. Specifically, they cannot algebraically determine whether a national surge is driven by compositional shifts in the population, such as increased healthcare access, or by changing behavioral propensities among medical providers [13,14]. Furthermore, the literature currently lacks an evaluation of the most recent demographic indicators, leaving the contemporary trajectory unquantified. Examining factors associated with exact percentage-point changes in prevalence over time becomes essential for designing targeted, sustainable public health regulations.

To the best of the author’s knowledge, no prior studies in Bangladesh have examined the determinants of the trends in Caesarean section deliveries using econometric decomposition methodologies. Therefore, this study aimed to investigate the factors associated with the change in Caesarean section prevalence in Bangladesh between 2012 and 2025. By applying a multivariate Linear Probability Model (LPM) Oaxaca-Blinder decomposition to pooled, nationally representative survey data, this study seeks to systematically separate the absolute contributions of population-level compositional shifts from behavioral and structural changes in obstetric practice.

## Materials and methods

### Data source, sampling procedure, and study variables

Data were analyzed from the 2012–2013, 2019, and 2024–2025 Bangladesh Multiple Indicator Cluster Surveys (MICS) to estimate the trends in Caesarean section deliveries [15–17]. The MICS, developed by UNICEF and implemented by the Bangladesh Bureau of Statistics (BBS), utilizes a rigorous two-stage stratified sampling design to generate nationally representative health and demographic estimates [18]. Census enumeration blocks act as the primary sampling units (PSUs), selected with probability proportional to size, followed by the systematic random selection of households within each PSU [15–18]. This study restricted the analysis to women of reproductive age (15–49 years) who delivered a child within the two years preceding the surveys to minimize recall bias [18,19]. After removing observations with missing data across the primary covariates, the sample size for each survey wave in the complete-case analytical cohort was 7,757 (2012), 9,271 (2019), and 9,506 (2025). Hence, the pooled descriptive sample across the three survey waves comprised 26,534 women. For the multivariate decomposition models, listwise deletion of observations with missing values across any covariate yielded a strict complete-case analytical cohort of N=16,179 for the primary 2012– 2025 macro comparison.

The outcome is delivery via Caesarean section, a binary variable coded ‘1’ if the delivery was performed via surgical intervention, and ‘0’ for vaginal delivery [2,6]. The independent variables were selected based on established epidemiological frameworks for maternal healthcare utilization and validated determinants of Caesarean section in Bangladesh and other low-and middle-income countries (LMICs) [6,12,20,21]. These included socio-demographic characteristics (maternal age at delivery, maternal education, household head education, wealth index quintile, type of residence, geographical region, religion of household head, sex of household head, child marriage), clinical and delivery characteristics (place of delivery, assistance at delivery, preceding birth interval, parity, previous child death, perceived baby size, pregnancy intent), maternal healthcare behaviors (skilled ANC provider, ANC visits) and media/empowerment indicators (mobile phone ownership, internet access) **(Table 1)**.

**Table 1:**
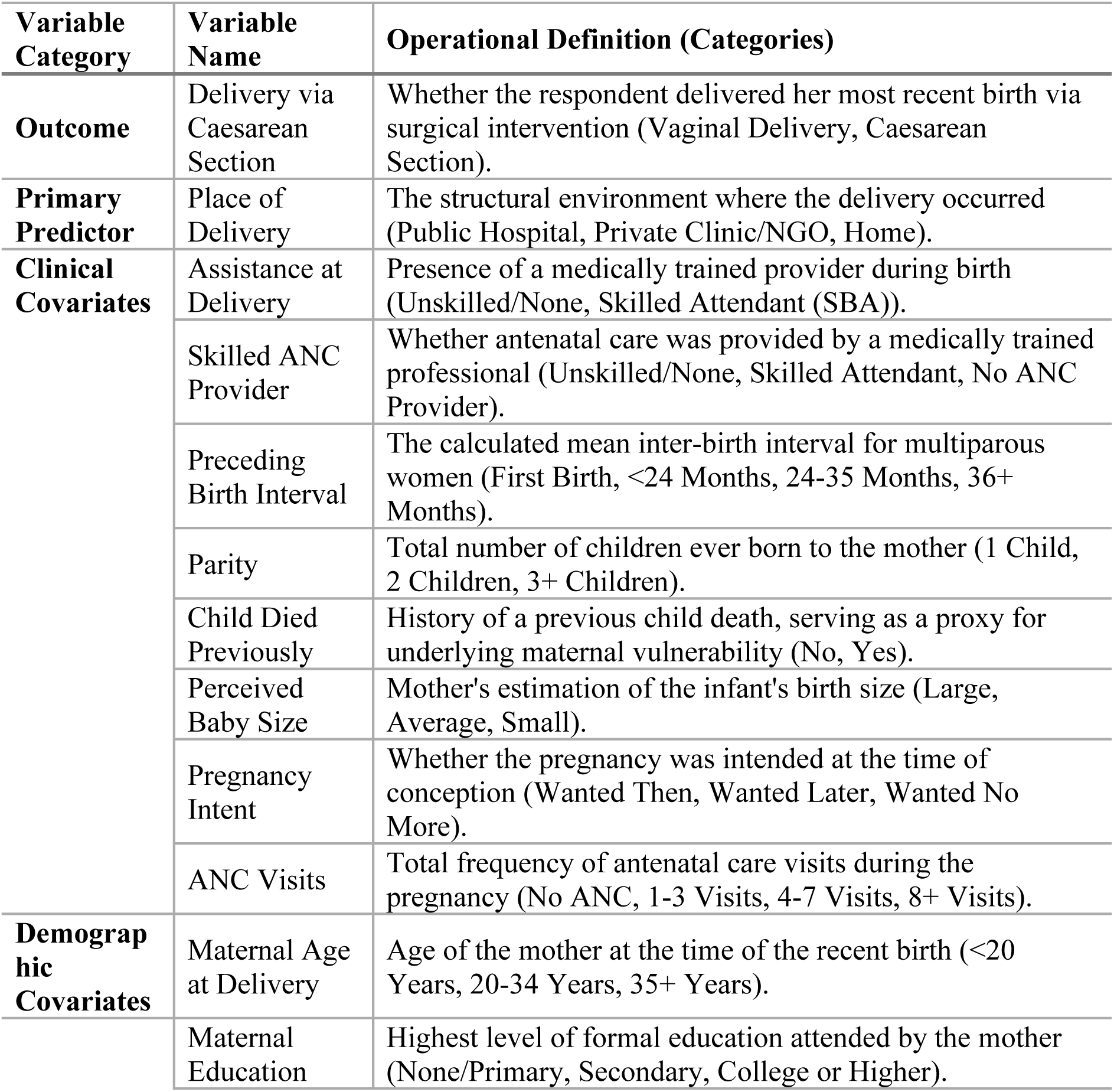
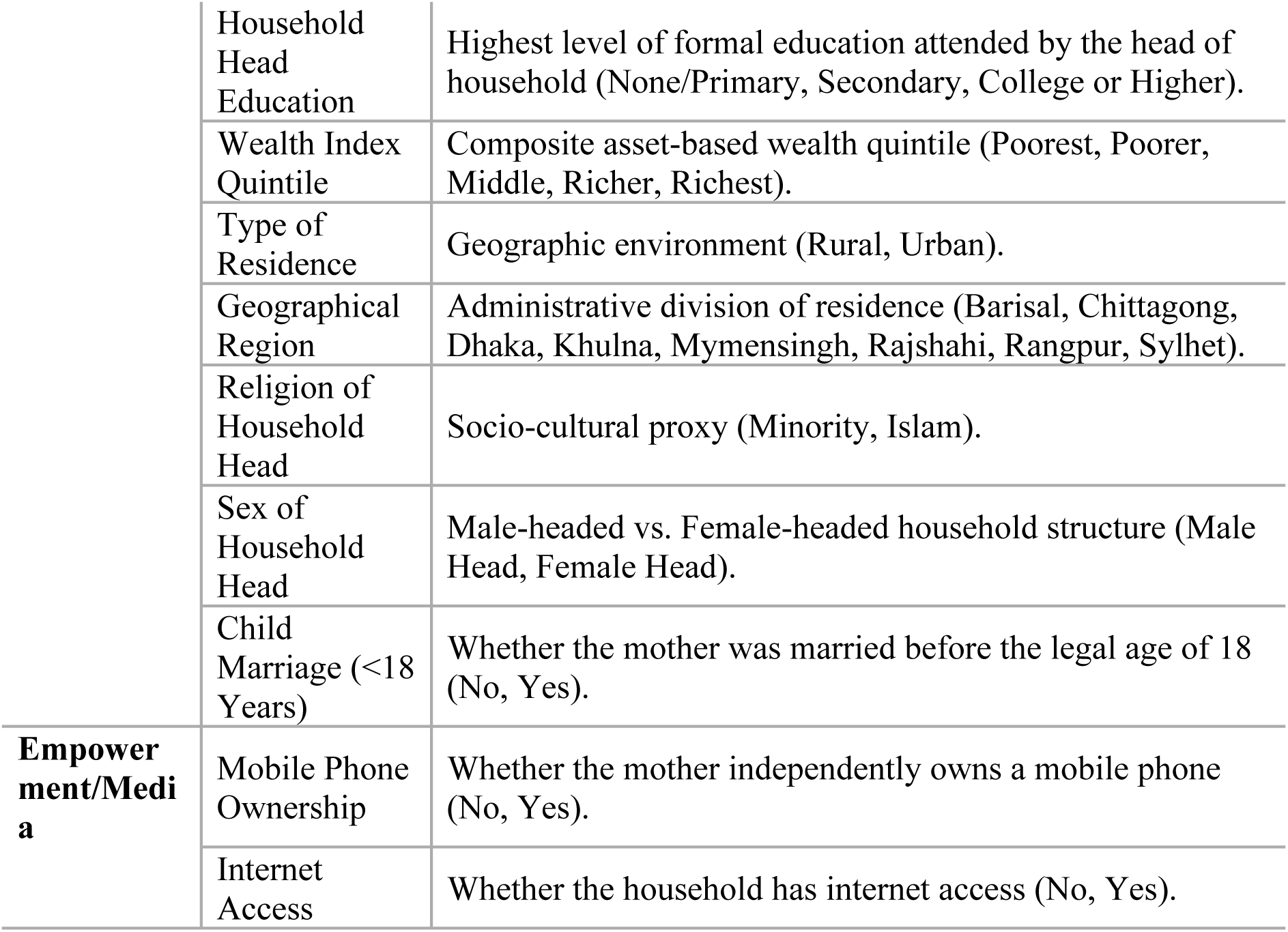
Description of the outcome and independent variables.

| Variable Category | Variable Name | Operational Definition (Categories) |
| --- | --- | --- |
| **Outcome** | Delivery via Caesarean Section | Whether the respondent delivered her most recent birth via surgical intervention (Vaginal Delivery, Caesarean Section). |
| **Primary Predictor** | Place of Delivery | The structural environment where the delivery occurred (Public Hospital, Private Clinic/NGO, Home). |
| **Clinical Covariates** | Assistance at Delivery | Presence of a medically trained provider during birth (Unskilled/None, Skilled Attendant (SBA)). |
|  | Skilled ANC Provider | Whether antenatal care was provided by a medically trained professional (Unskilled/None, Skilled Attendant, No ANC Provider). |
|  | Preceding Birth Interval | The calculated mean inter-birth interval for multiparous women (First Birth, <24 Months, 24-35 Months, 36+ Months). |
|  | Parity | Total number of children ever born to the mother (1 Child, 2 Children, 3+ Children). |
|  | Child Died Previously | History of a previous child death, serving as a proxy for underlying maternal vulnerability (No, Yes). |
|  | Perceived Baby Size | Mother's estimation of the infant's birth size (Large, Average, Small). |
|  | Pregnancy Intent | Whether the pregnancy was intended at the time of conception (Wanted Then, Wanted Later, Wanted No More). |
|  | ANC Visits | Total frequency of antenatal care visits during the pregnancy (No ANC, 1-3 Visits, 4-7 Visits, 8+ Visits). |
| **Demographic Covariates** | Maternal Age at Delivery | Age of the mother at the time of the recent birth (<20 Years, 20-34 Years, 35+ Years). |
|  | Maternal Education | Highest level of formal education attended by the mother (None/Primary, Secondary, College or Higher). |
|  | Household Head Education | Highest level of formal education attended by the head of household (None/Primary, Secondary, College or Higher). |
|  | Wealth Index Quintile | Composite asset-based wealth quintile (Poorest, Poorer, Middle, Richer, Richest). |
|  | Type of Residence | Geographic environment (Rural, Urban). |
|  | Geographical Region | Administrative division of residence (Barisal, Chittagong, Dhaka, Khulna, Mymensingh, Rajshahi, Rangpur, Sylhet). |
|  | Religion of Household Head | Socio-cultural proxy (Minority, Islam). |
|  | Sex of Household Head | Male-headed vs. Female-headed household structure (Male Head, Female Head). |
|  | Child Marriage (<18 Years) | Whether the mother was married before the legal age of 18 (No, Yes). |
| **Empowerment/Media** | Mobile Phone Ownership | Whether the mother independently owns a mobile phone (No, Yes). |
|  | Internet Access | Whether the household has internet access (No, Yes). |

To address systematic missingness inherent to survey skip-logic, strict data harmonization protocols were applied to antenatal care (ANC) variables. In the standard MICS architecture, women who report zero formal ANC visits bypass subsequent questions regarding provider type [15,17]. To prevent the artificial inflation of the medicalization baseline via listwise deletion, this skip-logic was recoded to establish distinct “No ANC Provider” and “No ANC Visits” categories, thereby retaining the highly vulnerable, unmedicalized cohorts within the decomposition models [14,22].

### Statistical analysis

All analyses were performed on weighted data using Stata to account for the representativeness and complex sampling procedures, employing the svy command suite [19]. Descriptive statistics for respondents’ characteristics were presented as percentages by survey year. The Caesarean section trends were examined separately for 2012–2019, 2019–2025, and the overall macro trajectory of 2012–2025. Bivariate distributional shifts across the three waves were evaluated using design-based Pearson (Rao-Scott) F-tests [19].

To quantify the determinants driving the observed temporal change in Caesarean section rates, a multivariate Oaxaca-Blinder decomposition model was utilized [23,24]. While decomposition is frequently applied to continuous outcomes or modeled via non-linear logistic functions [25], this study utilized a Linear Probability Model (LPM) framework for the binary outcome [26].

While Linear Probability Models (LPM) are theoretically susceptible to heteroskedasticity and out-of-bounds predictions, these limitations were structurally mitigated in this study. The application of Taylor-linearized variance estimation via the svy command inherently corrected for heteroskedasticity [19]. Furthermore, because the baseline and terminal outcome prevalences (19.48% to 51.95%) resided well within the intermediate probability range (20%– 80%), the linear approximation closely mimics the marginal effects of a non-linear logistic function, preventing the approximation decay associated with rare-event extremes [26,27]. The LPM approach guarantees exact additive decomposability of the components without the path-dependence and scaling limitations inherent to logit/probit decompositions, making it highly robust for structural policy analysis [26,27].

The change or difference in the proportion between two temporal cohorts can be mathematically attributed to differences in the composition between groups (endowments) and differences in the effects of independent variables (coefficients) [14]. The dependent variable in the LPM is a linear combination of covariates and regression coefficients:

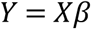

where *Y* is the *N* × 1 dependent variable vector, *X* is an *N* × *K* matrix of independent variables, and *β* is a *K* × 1 vector of coefficients [14]. The difference between group A (the recent survey) and group B (the baseline survey) in terms of the expected outcome *Y* can be decomposed into:

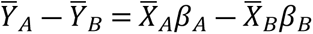

The difference in proportion in *Y* between groups A and B can be algebraically partitioned as:

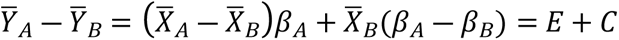

The component labeled **E** refers to the part of the difference attributable to changes in endowments or characteristics (compositional), usually called the explained component or characteristics effect [13,14]. The **C** component is the difference attributable to coefficients or behavioral change, which is usually labeled as the unexplained component [13,14]. Therefore, E represents a counterfactual comparison of the difference from the viewpoint of the data from the latest survey (e.g., 2025). It measures the expected absolute change in national Caesarean prevalence if the demographic distributions (endowments) of 2025 were applied to the baseline behavioral propensity (*β*) of 2012 [14]. C depicts a counterfactual comparison representing the anticipated difference if the respondents in the 2012 survey had experienced the behavioral reactions to Caesarean delivery seen among those in the 2025 survey [14].

To understand the contribution of each predictor, E and C were divided into portions or percentages of contribution (Pct.), *E_k_* and *C_k_* (*k* = 1,…,*k*; where K is the total number of independent variables) [13,14]. The expanded model utilized for this analysis was [14]:

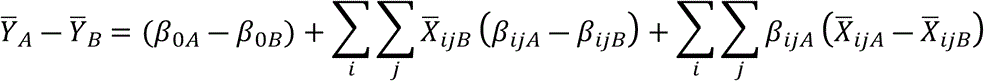

where:

- *β*_0*A*_ is the intercept in the regression equation for the recent survey (e.g., 2025)
- *β*_0*B*_ is the intercept in the regression equation for the baseline survey (e.g., 2012)
- *β_ijA_* is the coefficient of the *j^t^*^ℎ^ category of the *i^t^*^ℎ^ determinant for the recent survey
- *β_ijB_* is the coefficient of the *j^t^*^ℎ^ category of the *i^t^*^ℎ^ determinant for the baseline survey
- *X_ijA_* is the proportion of the *j^t^*^ℎ^ category of the *i^t^*^ℎ^ determinant for the recent survey
- *X_ijB_* is the proportion of the *j^t^*^ℎ^ category of the *i^t^*^ℎ^ determinant for the baseline survey

[14]

A macro decomposition model utilizing the strict complete-case cohort (N=16,179) was executed to evaluate the 2012–2025 trajectory. Sequential decompositions were subsequently run across the interim phases (2012–2019 and 2019–2025) to evaluate temporal fluctuations in the structural drivers. Statistical significance was established at *p* < 0.05.

### Ethical considerations

This study was a secondary analysis of fully anonymized, publicly available datasets from the UNICEF Global MICS Programme. The primary surveys were executed in accordance with the Declaration of Helsinki [28] and ethically cleared by the Bangladesh Bureau of Statistics and relevant national institutional review boards [15–17]. Written informed consent was obtained from all participants prior to primary data collection [15–17]. Formal authorization to access and analyze the microdata was granted by the UNICEF MICS platform; therefore, additional institutional ethical approval was not required.

Data were accessed for research purposes on September 12, 2026, and the author did not have access to information that could identify individual participants during or after data collection.

## Results

### Characteristics of the respondents

A total of 26,534 women who delivered a child within the two years preceding the surveys were included in the analysis (7,757 in 2012; 9,271 in 2019; and 9,506 in 2025). Across the 12-year study period, the demographic composition of the obstetric population experienced substantial shifts **(Table 2)**. The proportion of mothers who completed secondary education increased from 38.16% in 2012 to 54.19% in 2025. Concurrently, the proportion of mothers attaining college or higher education expanded substantially from 14.60% to 24.88%, alongside a notable decline in uneducated or primary-only mothers from 47.23% to 20.93% (*p* < 0.001). Simultaneously, maternal healthcare utilization expanded dramatically. Deliveries within health facilities surged by 39.78 percentage points, rising from 31.52% in 2012 to 71.30% in 2025. This medicalization was mirrored in antenatal care (ANC); the proportion of pregnancies receiving care from a skilled ANC provider increased from 55.98% in 2012 to 86.73% in 2025. Notably, the proportion of women receiving no ANC provider dropped precipitously from 35.52% in 2012 to just 8.12% in 2025 (*p* < 0.001), indicating near-universal integration of pregnant women into the formal healthcare system by the end of the study period.

**Table 2:**
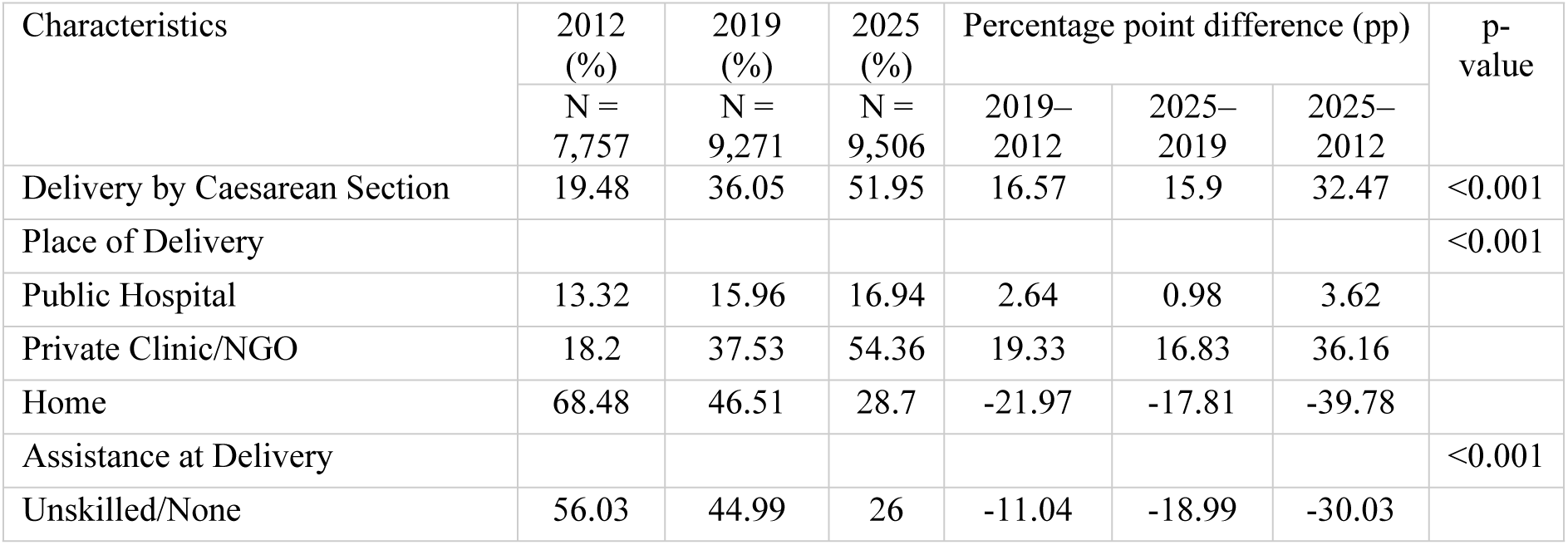
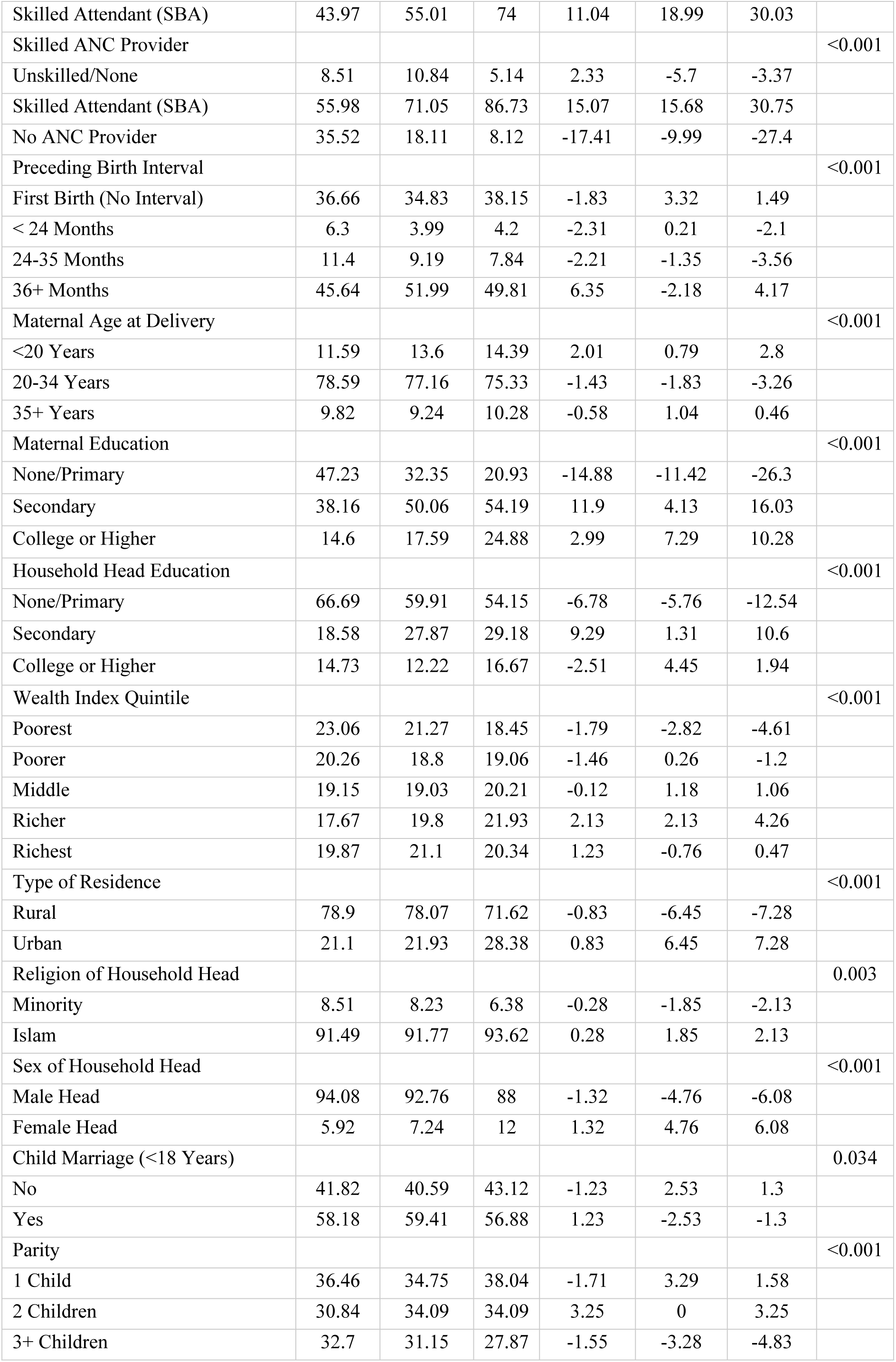
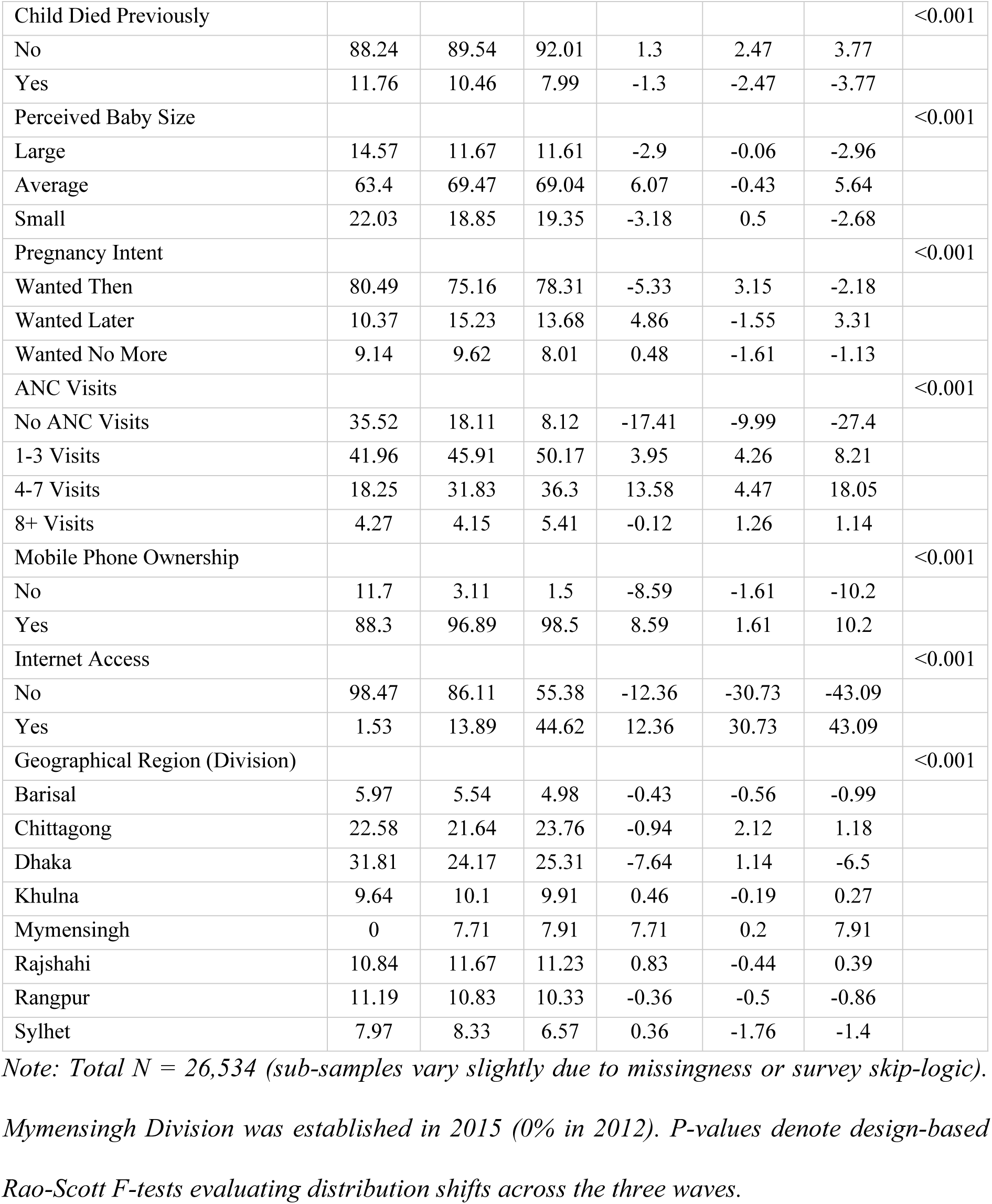
Percentage distribution of the respondents’ characteristics across the 2012, 2019, and 2025 Bangladesh MICS waves and the percentage point differences.

| Characteristics | 2012 (%) | 2019 (%) | 2025 (%) | Percentage point difference (pp) |  |  | p-value |
| --- | --- | --- | --- | --- | --- | --- | --- |
|  | N = 7,757 | N = 9,271 | N = 9,506 | 2019–2012 | 2025–2019 | 2025–2012 |  |
| Delivery by Caesarean Section | 19.48 | 36.05 | 51.95 | 16.57 | 15.9 | 32.47 | <0.001 |
| Place of Delivery |  |  |  |  |  |  | <0.001 |
| Public Hospital | 13.32 | 15.96 | 16.94 | 2.64 | 0.98 | 3.62 |  |
| Private Clinic/NGO | 18.2 | 37.53 | 54.36 | 19.33 | 16.83 | 36.16 |  |
| Home | 68.48 | 46.51 | 28.7 | -21.97 | -17.81 | -39.78 |  |
| Assistance at Delivery |  |  |  |  |  |  | <0.001 |
| Unskilled/None | 56.03 | 44.99 | 26 | -11.04 | -18.99 | -30.03 |  |
| Skilled Attendant (SBA) | 43.97 | 55.01 | 74 | 11.04 | 18.99 | 30.03 |  |
| Skilled ANC Provider |  |  |  |  |  |  | <0.001 |
| Unskilled/None | 8.51 | 10.84 | 5.14 | 2.33 | -5.7 | -3.37 |  |
| Skilled Attendant (SBA) | 55.98 | 71.05 | 86.73 | 15.07 | 15.68 | 30.75 |  |
| No ANC Provider | 35.52 | 18.11 | 8.12 | -17.41 | -9.99 | -27.4 |  |
| Preceding Birth Interval |  |  |  |  |  |  | <0.001 |
| First Birth (No Interval) | 36.66 | 34.83 | 38.15 | -1.83 | 3.32 | 1.49 |  |
| < 24 Months | 6.3 | 3.99 | 4.2 | -2.31 | 0.21 | -2.1 |  |
| 24-35 Months | 11.4 | 9.19 | 7.84 | -2.21 | -1.35 | -3.56 |  |
| 36+ Months | 45.64 | 51.99 | 49.81 | 6.35 | -2.18 | 4.17 |  |
| Maternal Age at Delivery |  |  |  |  |  |  | <0.001 |
| <20 Years | 11.59 | 13.6 | 14.39 | 2.01 | 0.79 | 2.8 |  |
| 20-34 Years | 78.59 | 77.16 | 75.33 | -1.43 | -1.83 | -3.26 |  |
| 35+ Years | 9.82 | 9.24 | 10.28 | -0.58 | 1.04 | 0.46 |  |
| Maternal Education |  |  |  |  |  |  | <0.001 |
| None/Primary | 47.23 | 32.35 | 20.93 | -14.88 | -11.42 | -26.3 |  |
| Secondary | 38.16 | 50.06 | 54.19 | 11.9 | 4.13 | 16.03 |  |
| College or Higher | 14.6 | 17.59 | 24.88 | 2.99 | 7.29 | 10.28 |  |
| Household Head Education |  |  |  |  |  |  | <0.001 |
| None/Primary | 66.69 | 59.91 | 54.15 | -6.78 | -5.76 | -12.54 |  |
| Secondary | 18.58 | 27.87 | 29.18 | 9.29 | 1.31 | 10.6 |  |
| College or Higher | 14.73 | 12.22 | 16.67 | -2.51 | 4.45 | 1.94 |  |
| Wealth Index Quintile |  |  |  |  |  |  | <0.001 |
| Poorest | 23.06 | 21.27 | 18.45 | -1.79 | -2.82 | -4.61 |  |
| Poorer | 20.26 | 18.8 | 19.06 | -1.46 | 0.26 | -1.2 |  |
| Middle | 19.15 | 19.03 | 20.21 | -0.12 | 1.18 | 1.06 |  |
| Richer | 17.67 | 19.8 | 21.93 | 2.13 | 2.13 | 4.26 |  |
| Richest | 19.87 | 21.1 | 20.34 | 1.23 | -0.76 | 0.47 |  |
| Type of Residence |  |  |  |  |  |  | <0.001 |
| Rural | 78.9 | 78.07 | 71.62 | -0.83 | -6.45 | -7.28 |  |
| Urban | 21.1 | 21.93 | 28.38 | 0.83 | 6.45 | 7.28 |  |
| Religion of Household Head |  |  |  |  |  |  | 0.003 |
| Minority | 8.51 | 8.23 | 6.38 | -0.28 | -1.85 | -2.13 |  |
| Islam | 91.49 | 91.77 | 93.62 | 0.28 | 1.85 | 2.13 |  |
| Sex of Household Head |  |  |  |  |  |  | <0.001 |
| Male Head | 94.08 | 92.76 | 88 | -1.32 | -4.76 | -6.08 |  |
| Female Head | 5.92 | 7.24 | 12 | 1.32 | 4.76 | 6.08 |  |
| Child Marriage (<18 Years) |  |  |  |  |  |  | 0.034 |
| No | 41.82 | 40.59 | 43.12 | -1.23 | 2.53 | 1.3 |  |
| Yes | 58.18 | 59.41 | 56.88 | 1.23 | -2.53 | -1.3 |  |
| Parity |  |  |  |  |  |  | <0.001 |
| 1 Child | 36.46 | 34.75 | 38.04 | -1.71 | 3.29 | 1.58 |  |
| 2 Children | 30.84 | 34.09 | 34.09 | 3.25 | 0 | 3.25 |  |
| 3+ Children | 32.7 | 31.15 | 27.87 | -1.55 | -3.28 | -4.83 |  |
| Child Died Previously |  |  |  |  |  |  | <0.001 |
| No | 88.24 | 89.54 | 92.01 | 1.3 | 2.47 | 3.77 |  |
| Yes | 11.76 | 10.46 | 7.99 | -1.3 | -2.47 | -3.77 |  |
| Perceived Baby Size |  |  |  |  |  |  | <0.001 |
| Large | 14.57 | 11.67 | 11.61 | -2.9 | -0.06 | -2.96 |  |
| Average | 63.4 | 69.47 | 69.04 | 6.07 | -0.43 | 5.64 |  |
| Small | 22.03 | 18.85 | 19.35 | -3.18 | 0.5 | -2.68 |  |
| Pregnancy Intent |  |  |  |  |  |  | <0.001 |
| Wanted Then | 80.49 | 75.16 | 78.31 | -5.33 | 3.15 | -2.18 |  |
| Wanted Later | 10.37 | 15.23 | 13.68 | 4.86 | -1.55 | 3.31 |  |
| Wanted No More | 9.14 | 9.62 | 8.01 | 0.48 | -1.61 | -1.13 |  |
| ANC Visits |  |  |  |  |  |  | <0.001 |
| No ANC Visits | 35.52 | 18.11 | 8.12 | -17.41 | -9.99 | -27.4 |  |
| 1-3 Visits | 41.96 | 45.91 | 50.17 | 3.95 | 4.26 | 8.21 |  |
| 4-7 Visits | 18.25 | 31.83 | 36.3 | 13.58 | 4.47 | 18.05 |  |
| 8+ Visits | 4.27 | 4.15 | 5.41 | -0.12 | 1.26 | 1.14 |  |
| Mobile Phone Ownership |  |  |  |  |  |  | <0.001 |
| No | 11.7 | 3.11 | 1.5 | -8.59 | -1.61 | -10.2 |  |
| Yes | 88.3 | 96.89 | 98.5 | 8.59 | 1.61 | 10.2 |  |
| Internet Access |  |  |  |  |  |  | <0.001 |
| No | 98.47 | 86.11 | 55.38 | -12.36 | -30.73 | -43.09 |  |
| Yes | 1.53 | 13.89 | 44.62 | 12.36 | 30.73 | 43.09 |  |
| Geographical Region (Division) |  |  |  |  |  |  | <0.001 |
| Barisal | 5.97 | 5.54 | 4.98 | -0.43 | -0.56 | -0.99 |  |
| Chittagong | 22.58 | 21.64 | 23.76 | -0.94 | 2.12 | 1.18 |  |
| Dhaka | 31.81 | 24.17 | 25.31 | -7.64 | 1.14 | -6.5 |  |
| Khulna | 9.64 | 10.1 | 9.91 | 0.46 | -0.19 | 0.27 |  |
| Mymensingh | 0 | 7.71 | 7.91 | 7.71 | 0.2 | 7.91 |  |
| Rajshahi | 10.84 | 11.67 | 11.23 | 0.83 | -0.44 | 0.39 |  |
| Rangpur | 11.19 | 10.83 | 10.33 | -0.36 | -0.5 | -0.86 |  |
| Sylhet | 7.97 | 8.33 | 6.57 | 0.36 | -1.76 | -1.4 |  |

*Note: Total N = 26,534 (sub-samples vary slightly due to missingness or survey skip-logic).*
*Mymensingh Division was established in 2015 (0% in 2012). P-values denote design-based*
*Rao-Scott F-tests evaluating distribution shifts across the three waves.*

### Trends in Caesarean section delivery

The national prevalence of Caesarean section deliveries surged by 32.47 percentage points over the 12-year period, rising from 19.48% in 2012 to 36.05% in 2019, and reaching 51.95% by 2025 (*p* < 0.001) **(Figure 1)**.

**Figure 1:**
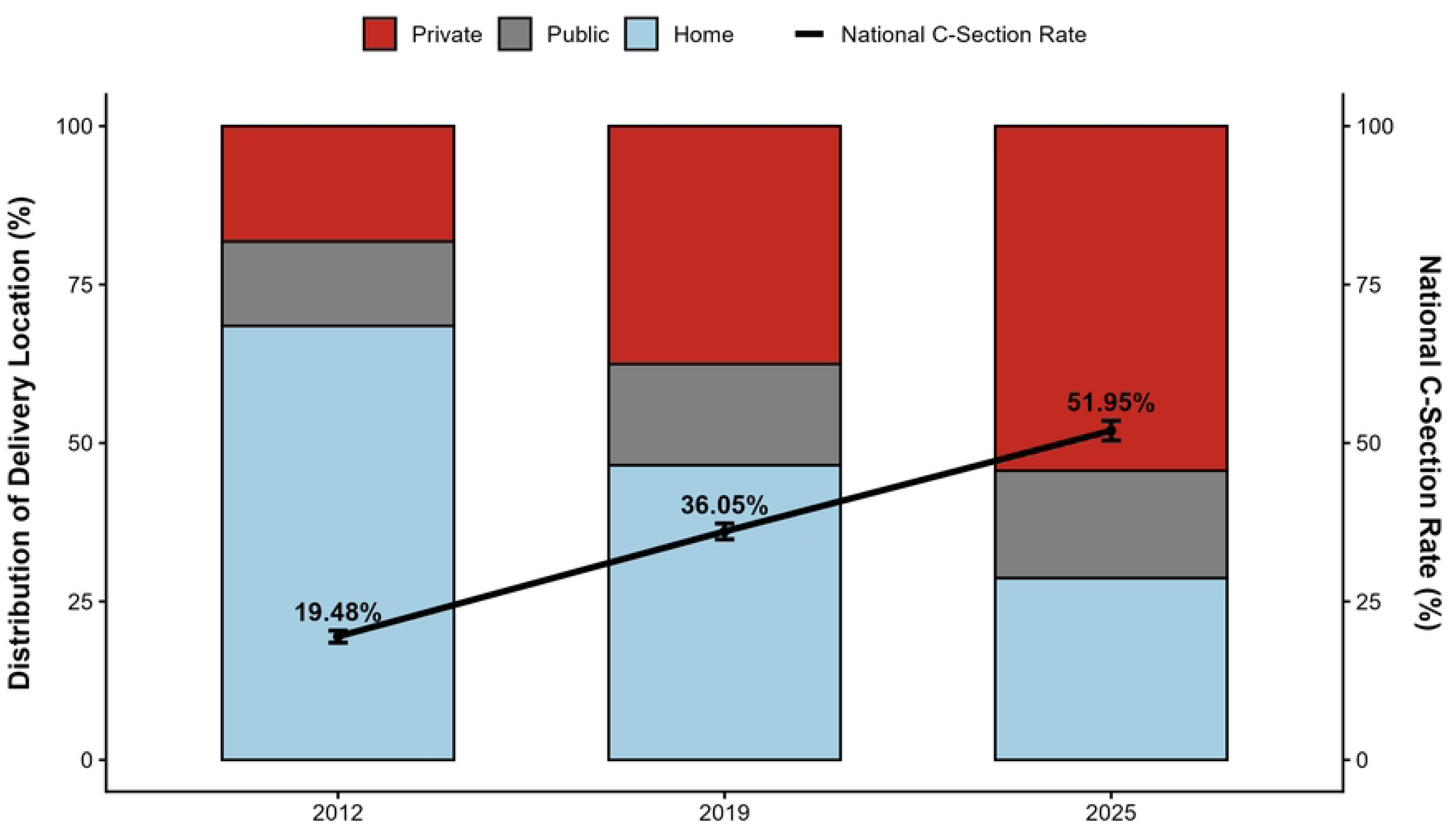
Trends in delivery location and national Caesarean section rates in Bangladesh, 2012–2025.

This increase was disproportionately concentrated within the private healthcare sector **(Figure 2)**. While Caesarean deliveries within public hospitals experienced a slight decline (from 38.66% in 2012 to 33.51% in 2025), the rate within private clinics/NGOs remained exceptionally high and continued to climb, rising from 78.73% in 2012 to 85.12% in 2025.

**Figure 2:**
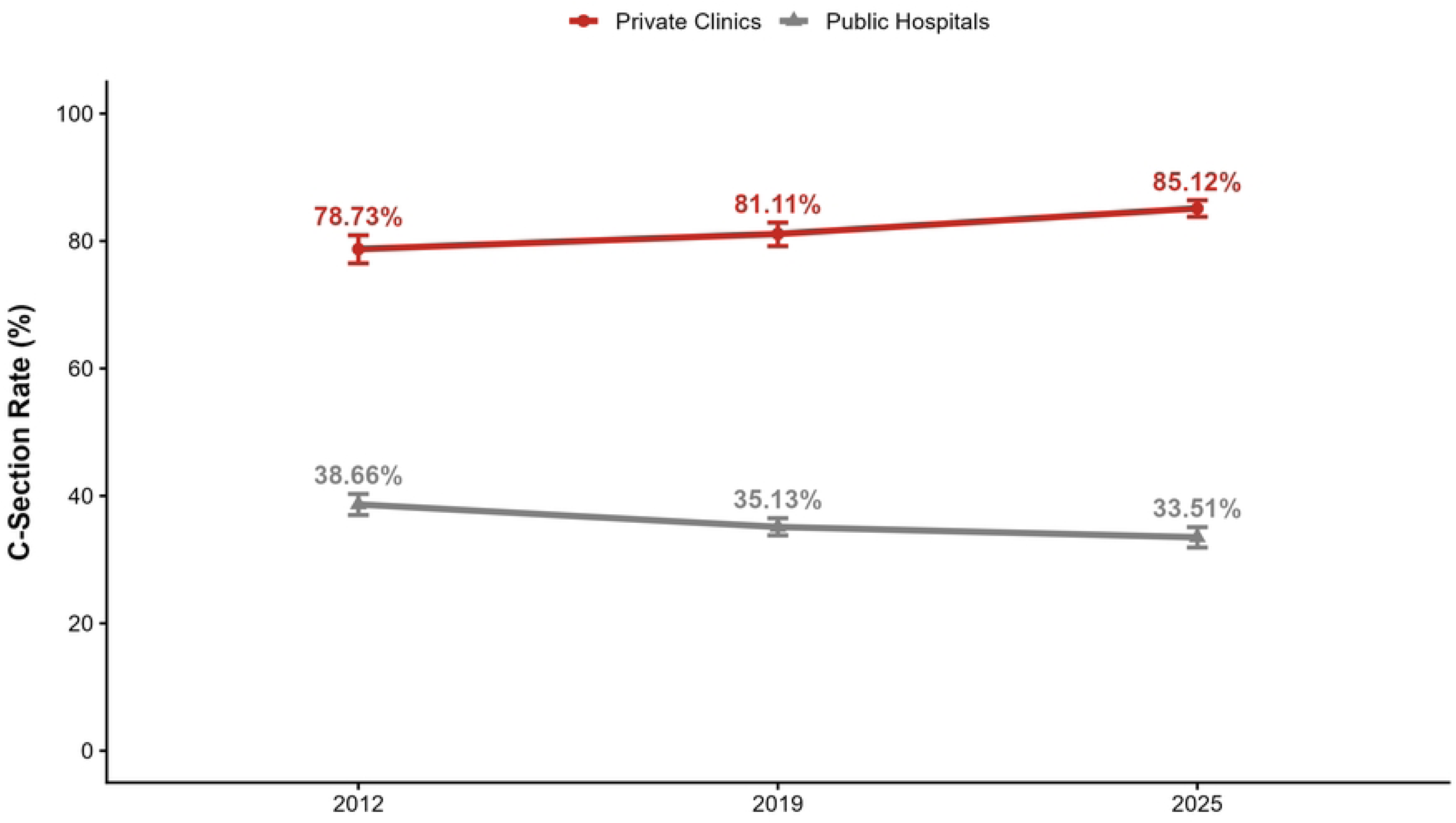
Caesarean section rates by institutional delivery setting in Bangladesh, 2012–2025.

Simultaneously, the integration of previously unmedicalized populations into the clinical pathway exposed them to higher surgical rates **(Table 3)**. Among women reporting zero ANC visits, the Caesarean section rate was only 3.96% in 2012. However, among women achieving 8 or more ANC visits, the surgical rate reached 48.34% in 2012 and escalated to 78.99% by 2025. Across virtually all maternal age groups, wealth index quintiles, and geographical regions, the likelihood of surgical delivery increased significantly between 2012 and 2025.

**Table 3:** Trends in the percentage of Caesarean section deliveries by respondents’ characteristics across 2012, 2019, and 2025.

| Characteristics | 2012 (%) | 2019 (%) | 2025 (%) | Percentage point difference (pp) |  |  | p-value |
| --- | --- | --- | --- | --- | --- | --- | --- |
|  | N = 7,757 | N = 9,271 | N = 9,506 | 2019–2012 | 2025–2019 | 2025–2012 |  |
| Place of Delivery |  |  |  |  |  |  | <0.001 |
| Public Hospital | 38.66 | 35.13 | 33.51 | -3.53 | -1.62 | -5.15 |  |
| Private Clinic/NGO | 78.73 | 81.11 | 85.12 | 2.38 | 4.01 | 6.39 |  |
| Home | 0 | 0 | 0 | 0 | 0 | 0 |  |
| Assistance at Delivery |  |  |  |  |  |  | <0.001 |
| Unskilled/None | 0.23 | 0.14 | 1.34 | -0.09 | 1.2 | 1.11 |  |
| Skilled Attendant (SBA) | 44.01 | 65.42 | 69.73 | 21.41 | 4.31 | 25.72 |  |
| Skilled ANC Provider |  |  |  |  |  |  | <0.001 |
| Unskilled/None | 8.18 | 17.31 | 30.47 | 9.13 | 13.16 | 22.29 |  |
| Skilled Attendant (SBA) | 27.13 | 44.31 | 55.17 | 17.18 | 10.86 | 28.04 |  |
| No ANC Provider | 3.96 | 9.59 | 17.23 | 5.63 | 7.64 | 13.27 |  |
| Preceding Birth Interval |  |  |  |  |  |  | <0.001 |
| First Birth (No Interval) | 28 | 44.54 | 58.69 | 16.54 | 14.15 | 30.69 |  |
| < 24 Months | 17.24 | 32.65 | 51.01 | 15.41 | 18.36 | 33.77 |  |
| 24-35 Months | 7.3 | 25.64 | 41.22 | 18.34 | 15.58 | 33.92 |  |
| 36+ Months | 16.19 | 32.53 | 48.8 | 16.34 | 16.27 | 32.61 |  |
| Maternal Age at Delivery |  |  |  |  |  |  | <0.001 |
| <20 Years | 19.33 | 34.97 | 48.81 | 15.64 | 13.84 | 29.48 |  |
| 20-34 Years | 20.44 | 37.45 | 52.89 | 17.01 | 15.44 | 32.45 |  |
| 35+ Years | 11.96 | 25.93 | 49.42 | 13.97 | 23.49 | 37.46 |  |
| Maternal Education |  |  |  |  |  |  | <0.001 |
| None/Primary | 8.19 | 18.63 | 33.53 | 10.44 | 14.9 | 25.34 |  |
| Secondary | 22.38 | 37.96 | 50.99 | 15.58 | 13.03 | 28.61 |  |
| College or Higher | 48.44 | 62.64 | 69.5 | 14.2 | 6.86 | 21.06 |  |
| Household Head Education |  |  |  |  |  |  | <0.001 |
| None/Primary | 11.86 | 27.15 | 44.67 | 15.29 | 17.52 | 32.81 |  |
| Secondary | 24.96 | 44.28 | 56.28 | 19.32 | 12 | 31.32 |  |
| College or Higher | 47.11 | 60.9 | 68.01 | 13.79 | 7.11 | 20.9 |  |
| Wealth Index Quintile |  |  |  |  |  |  | <0.001 |
| Poorest | 5.53 | 13.19 | 34.37 | 7.66 | 21.18 | 28.84 |  |
| Poorer | 8.81 | 26.09 | 44.97 | 17.28 | 18.88 | 36.16 |  |
| Middle | 13 | 36.54 | 52.24 | 23.54 | 15.7 | 39.24 |  |
| Richer | 25.78 | 44.55 | 57.36 | 18.77 | 12.81 | 31.58 |  |
| Richest | 47.19 | 59.55 | 68.29 | 12.36 | 8.74 | 21.1 |  |
| Type of Residence |  |  |  |  |  |  | <0.001 |
| Rural | 15.66 | 32.86 | 50.49 | 17.2 | 17.63 | 34.83 |  |
| Urban | 33.78 | 47.4 | 55.63 | 13.62 | 8.23 | 21.85 |  |
| Religion of Household Head |  |  |  |  |  |  | <0.001 |
| Minority | 25.62 | 43.86 | 64.7 | 18.24 | 20.84 | 39.08 |  |
| Islam | 18.91 | 35.35 | 51.08 | 16.44 | 15.73 | 32.17 |  |
| Sex of Household Head |  |  |  |  |  |  | 0.002 |
| Male Head | 19.07 | 35.67 | 51.14 | 16.6 | 15.47 | 32.07 |  |
| Female Head | 25.96 | 40.87 | 57.86 | 14.91 | 16.99 | 31.9 |  |
| Child Marriage (<18 Years) |  |  |  |  |  |  | <0.001 |
| No | 24.8 | 40.77 | 56.93 | 15.97 | 16.16 | 32.13 |  |
| Yes | 15.66 | 32.81 | 48.16 | 17.15 | 15.35 | 32.5 |  |
| Parity |  |  |  |  |  |  | <0.001 |
| 1 Child | 28 | 44.54 | 58.69 | 16.54 | 14.15 | 30.69 |  |
| 2 Children | 20.58 | 38.54 | 54.63 | 17.96 | 16.09 | 34.05 |  |
| 3+ Children | 8.95 | 23.86 | 39.46 | 14.91 | 15.6 | 30.51 |  |
| Child Died Previously |  |  |  |  |  |  | <0.001 |
| No | 20.65 | 36.74 | 53.03 | 16.09 | 16.29 | 32.38 |  |
| Yes | 10.74 | 30.17 | 39.49 | 19.43 | 9.32 | 28.75 |  |
| Perceived Baby Size |  |  |  |  |  |  | <0.001 |
| Large | 31.5 | 47.73 | 51.84 | 16.23 | 4.11 | 20.34 |  |
| Average | 18.93 | 34.85 | 52.77 | 15.92 | 17.92 | 33.84 |  |
| Small | 16.46 | 34.05 | 50.28 | 17.59 | 16.23 | 33.82 |  |
| Pregnancy Intent |  |  |  |  |  |  | <0.001 |
| Wanted Then | 20.57 | 37.92 | 53.49 | 17.35 | 15.57 | 32.92 |  |
| Wanted Later | 20.64 | 34.01 | 50.38 | 13.37 | 16.37 | 29.74 |  |
| Wanted No More | 8.67 | 24.66 | 39.49 | 15.99 | 14.83 | 30.82 |  |
| ANC Visits |  |  |  |  |  |  | <0.001 |
| No ANC Visits | 3.96 | 9.59 | 17.23 | 5.63 | 7.64 | 13.27 |  |
| 1-3 Visits | 18.71 | 29.86 | 42.67 | 11.15 | 12.81 | 23.96 |  |
| 4-7 Visits | 32.7 | 53 | 65.4 | 20.3 | 12.4 | 32.7 |  |
| 8+ Visits | 48.34 | 67.01 | 78.99 | 18.67 | 11.98 | 30.65 |  |
| Mobile Phone Ownership |  |  |  |  |  |  | <0.001 |
| No | 6.03 | 25.35 | 42.12 | 19.32 | 16.77 | 36.09 |  |
| Yes | 21.27 | 36.39 | 52.09 | 15.12 | 15.7 | 30.82 |  |
| Internet Access |  |  |  |  |  |  | <0.001 |
| No | 18.75 | 32.35 | 45.45 | 13.6 | 13.1 | 26.7 |  |
| Yes | 80.48 | 57.63 | 54.08 | -22.85 | -3.55 | -26.4 |  |
| Geographical Region (Division) |  |  |  |  |  |  | <0.001 |
| Barisal | 10.66 | 25.9 | 45.76 | 15.24 | 19.86 | 35.1 |  |
| Chittagong | 15.22 | 29.41 | 40.39 | 14.19 | 10.98 | 25.17 |  |
| Dhaka | 24.53 | 46.92 | 60.23 | 22.39 | 13.31 | 35.7 |  |
| Khulna | 30.82 | 52.62 | 71.2 | 21.8 | 18.58 | 40.38 |  |
| Mymensingh | N/A | 22.02 | 43.95 | N/A | 21.93 | N/A |  |
| Rajshahi | 22.52 | 40.69 | 59.65 | 18.17 | 18.96 | 37.13 |  |
| Rangpur | 11.84 | 32.29 | 51.08 | 20.45 | 18.79 | 39.24 |  |
| Sylhet | 10.88 | 19.78 | 35.32 | 8.9 | 15.54 | 24.44 |  |

*The p-value represents the design-based Pearson (Rao-Scott) F-test for the cross-tabulation of the Caesarean section outcome across all survey waves. Mymensingh Division was established in 2015, resulting in an N/A for 2012 baseline trend arithmetic. 'No ANC Provider' and 'No ANC Visits' (as well as 'First Birth' and 'Parity: 1 Child') represent the same underlying unmedicalized and primiparous sub-cohorts across their respective variable classifications.*

Furthermore, this surgical proliferation rapidly expanded into previously low-intervention demographics. The Caesarean section rate among the poorest wealth quintile multiplied more than sixfold, jumping 28.84 percentage points from 5.53% in 2012 to 34.37% in 2025. A similarly stark geographical homogenization occurred, with the surgical rate among rural women surging by 34.83 percentage points (from 15.66% to 50.49%), rapidly closing the historical gap with urban populations.

### Factors associated with the surge in Caesarean sections

To quantify the determinants driving the 32.69 percentage-point macro surge in the complete-case analytical cohort (N=16,179) between 2012 and 2025, a multivariate Oaxaca-Blinder decomposition was performed **(Table 4)**. The analysis reveals that the epidemic is overwhelmingly driven by differences in compositional factors (endowments) rather than differences in behavioral responses (coefficients). At the aggregate level, compositional shifts in the obstetric population explained 87.77% of the overall national increase (Coefficient: -0.2869; 95% CI: -0.3096, -0.2641; p<0.001), while behavioral factors explained only 12.23% of the variance (Coefficient: -0.0400; 95% CI: -0.0638, -0.0162; p=0.001).

**Table 4.**
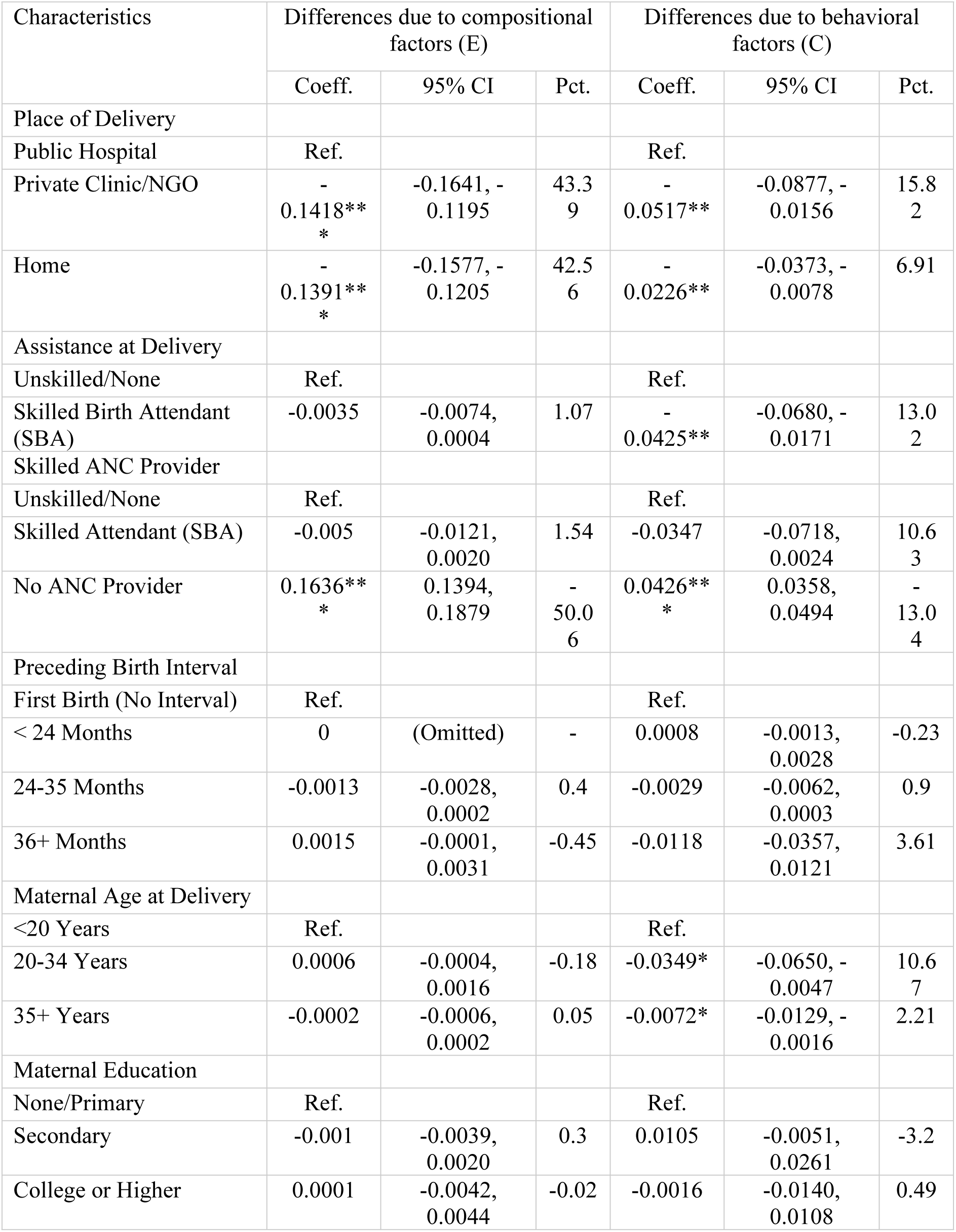
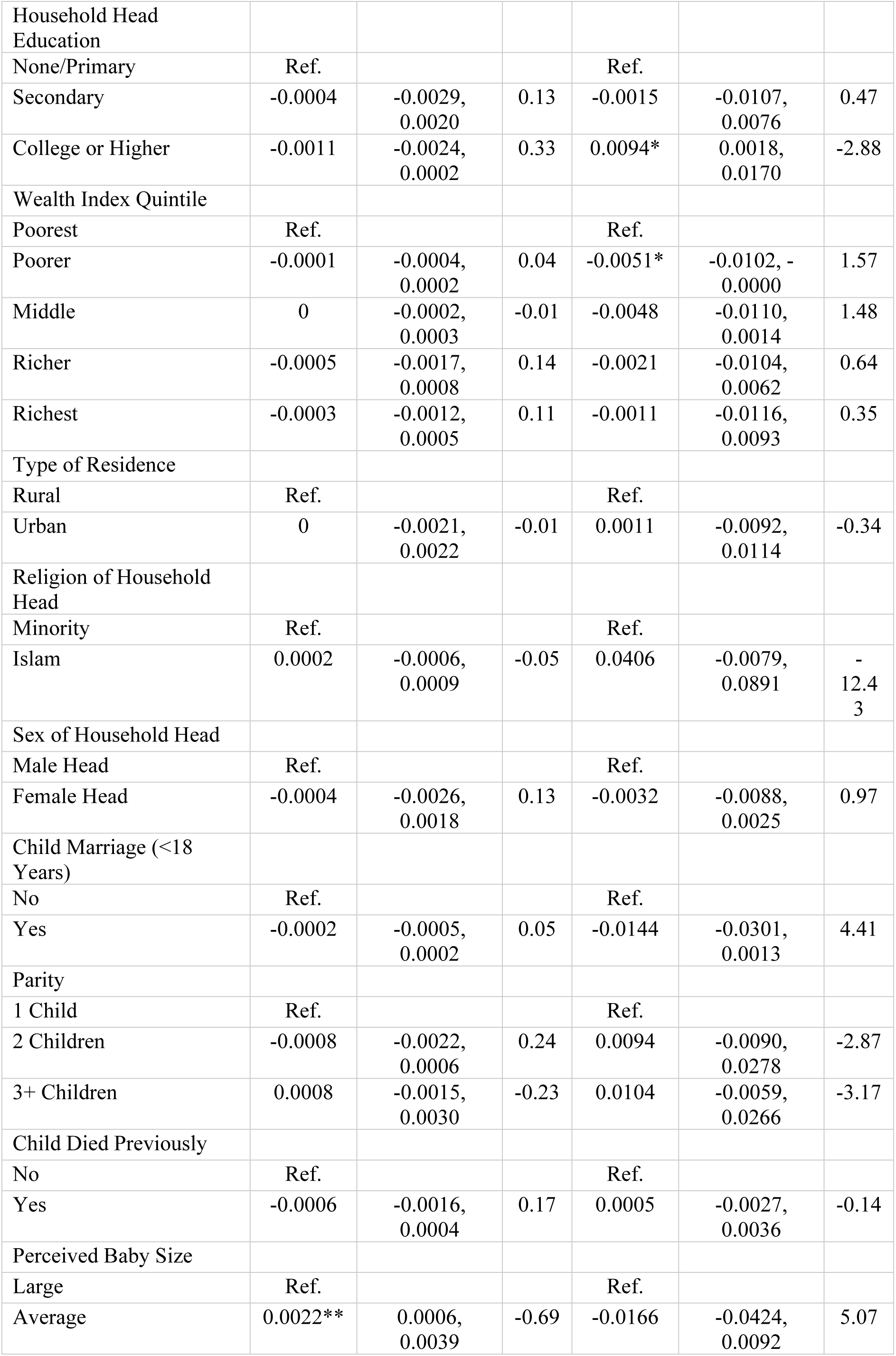
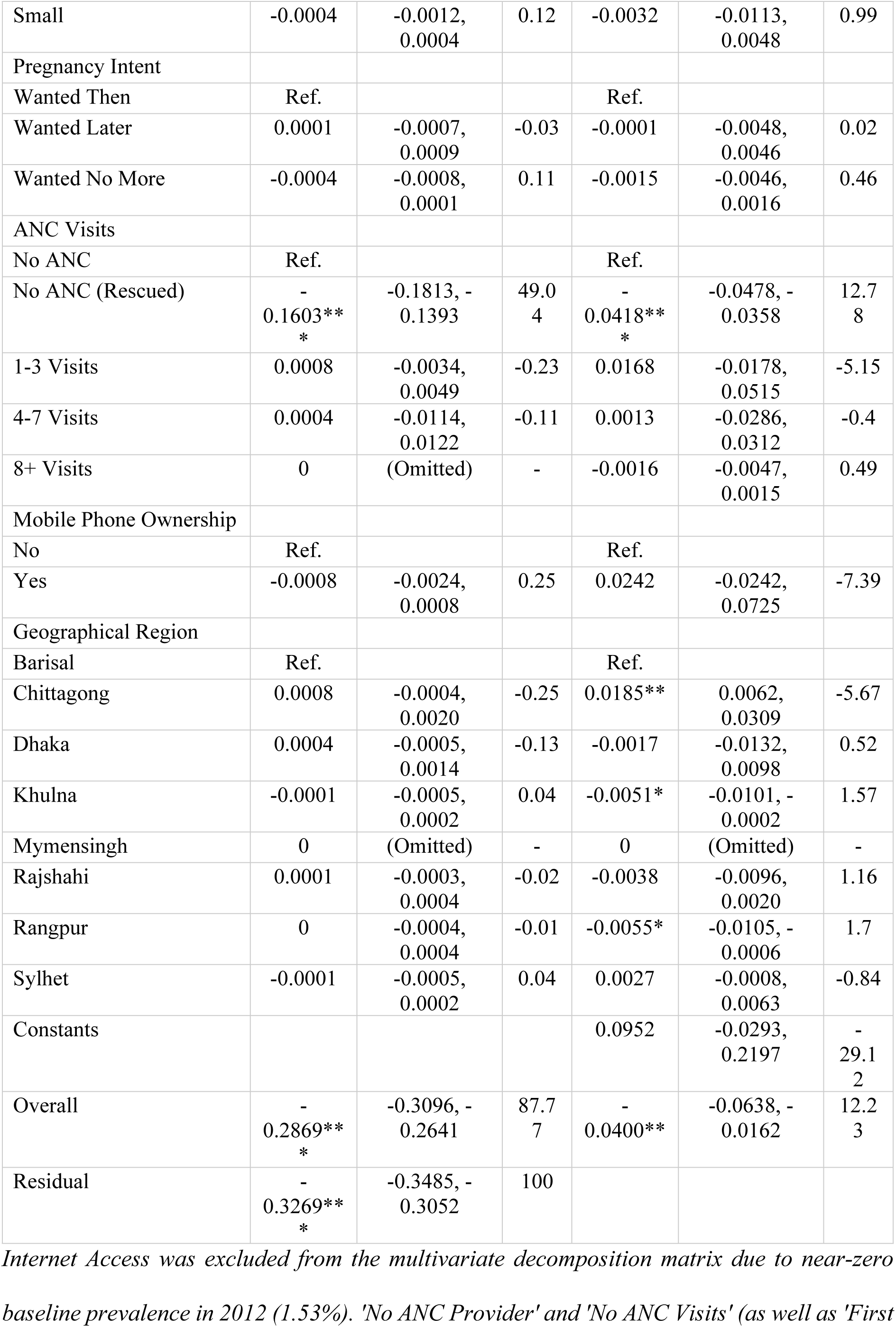
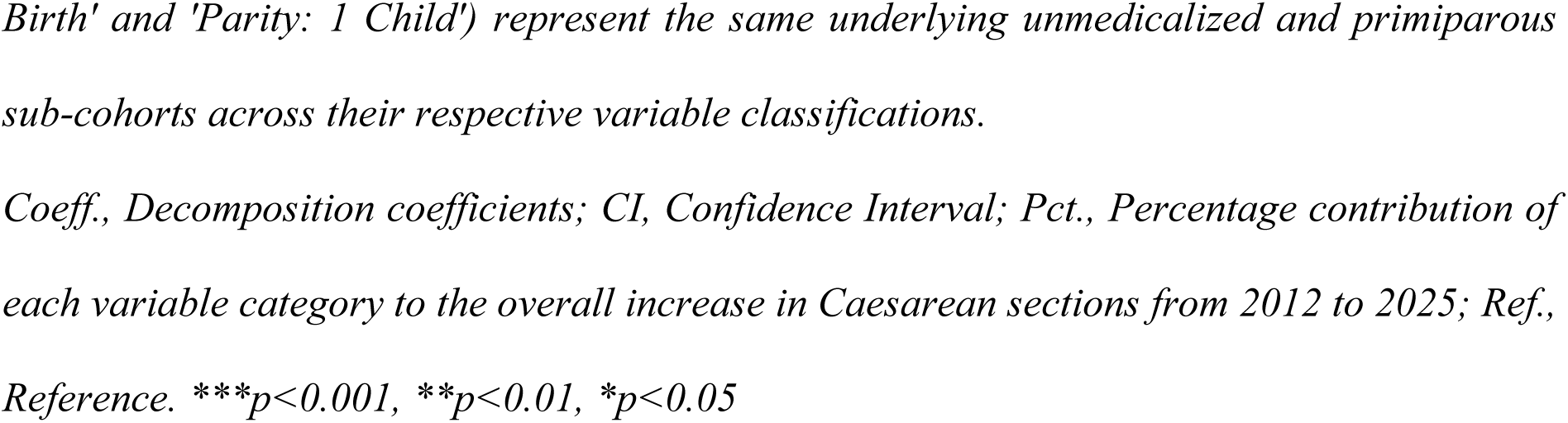
Multivariate Oaxaca-Blinder Decomposition analysis for the change in the percentage of Caesarean sections in Bangladesh (2012–2025).

| Characteristics | Differences due to compositional factors (E) |  |  | Differences due to behavioral factors (C) |  |  |
| --- | --- | --- | --- | --- | --- | --- |
|  | Coeff. | 95% CI | Pct. | Coeff. | 95% CI | Pct. |
| Place of Delivery |  |  |  |  |  |  |
| Public Hospital | Ref. |  |  | Ref. |  |  |
| Private Clinic/NGO | -0.1418**<br>* | -0.1641, -0.1195 | 43.3<br>9 | -0.0517** | -0.0877, -0.0156 | 15.8<br>2 |
| Home | -0.1391**<br>* | -0.1577, -0.1205 | 42.5<br>6 | -0.0226** | -0.0373, -0.0078 | 6.91 |
| Assistance at Delivery |  |  |  |  |  |  |
| Unskilled/None | Ref. |  |  | Ref. |  |  |
| Skilled Birth Attendant (SBA) | -0.0035 | -0.0074, 0.0004 | 1.07 | -0.0425** | -0.0680, -0.0171 | 13.0<br>2 |
| Skilled ANC Provider |  |  |  |  |  |  |
| Unskilled/None | Ref. |  |  | Ref. |  |  |
| Skilled Attendant (SBA) | -0.005 | -0.0121, 0.0020 | 1.54 | -0.0347 | -0.0718, 0.0024 | 10.6<br>3 |
| No ANC Provider | 0.1636**<br>* | 0.1394, 0.1879 | -50.0<br>6 | 0.0426**<br>* | 0.0358, 0.0494 | -13.0<br>4 |
| Preceding Birth Interval |  |  |  |  |  |  |
| First Birth (No Interval) | Ref. |  |  | Ref. |  |  |
| < 24 Months | 0 | (Omitted) | - | 0.0008 | -0.0013, 0.0028 | -0.23 |
| 24-35 Months | -0.0013 | -0.0028, 0.0002 | 0.4 | -0.0029 | -0.0062, 0.0003 | 0.9 |
| 36+ Months | 0.0015 | -0.0001, 0.0031 | -0.45 | -0.0118 | -0.0357, 0.0121 | 3.61 |
| Maternal Age at Delivery |  |  |  |  |  |  |
| <20 Years | Ref. |  |  | Ref. |  |  |
| 20-34 Years | 0.0006 | -0.0004, 0.0016 | -0.18 | -0.0349* | -0.0650, -0.0047 | 10.6<br>7 |
| 35+ Years | -0.0002 | -0.0006, 0.0002 | 0.05 | -0.0072* | -0.0129, -0.0016 | 2.21 |
| Maternal Education |  |  |  |  |  |  |
| None/Primary | Ref. |  |  | Ref. |  |  |
| Secondary | -0.001 | -0.0039, 0.0020 | 0.3 | 0.0105 | -0.0051, 0.0261 | -3.2 |
| College or Higher | 0.0001 | -0.0042, 0.0044 | -0.02 | -0.0016 | -0.0140, 0.0108 | 0.49 |
| Household Head Education |  |  |  |  |  |  |
| None/Primary | Ref. |  |  | Ref. |  |  |
| Secondary | -0.0004 | -0.0029,<br>0.0020 | 0.13 | -0.0015 | -0.0107,<br>0.0076 | 0.47 |
| College or Higher | -0.0011 | -0.0024,<br>0.0002 | 0.33 | 0.0094* | 0.0018,<br>0.0170 | -2.88 |
| Wealth Index Quintile |  |  |  |  |  |  |
| Poorest | Ref. |  |  | Ref. |  |  |
| Poorer | -0.0001 | -0.0004,<br>0.0002 | 0.04 | -0.0051* | -0.0102, -<br>0.0000 | 1.57 |
| Middle | 0 | -0.0002,<br>0.0003 | -0.01 | -0.0048 | -0.0110,<br>0.0014 | 1.48 |
| Richer | -0.0005 | -0.0017,<br>0.0008 | 0.14 | -0.0021 | -0.0104,<br>0.0062 | 0.64 |
| Richest | -0.0003 | -0.0012,<br>0.0005 | 0.11 | -0.0011 | -0.0116,<br>0.0093 | 0.35 |
| Type of Residence |  |  |  |  |  |  |
| Rural | Ref. |  |  | Ref. |  |  |
| Urban | 0 | -0.0021,<br>0.0022 | -0.01 | 0.0011 | -0.0092,<br>0.0114 | -0.34 |
| Religion of Household Head |  |  |  |  |  |  |
| Minority | Ref. |  |  | Ref. |  |  |
| Islam | 0.0002 | -0.0006,<br>0.0009 | -0.05 | 0.0406 | -0.0079,<br>0.0891 | -<br>12.4<br>3 |
| Sex of Household Head |  |  |  |  |  |  |
| Male Head | Ref. |  |  | Ref. |  |  |
| Female Head | -0.0004 | -0.0026,<br>0.0018 | 0.13 | -0.0032 | -0.0088,<br>0.0025 | 0.97 |
| Child Marriage (<18 Years) |  |  |  |  |  |  |
| No | Ref. |  |  | Ref. |  |  |
| Yes | -0.0002 | -0.0005,<br>0.0002 | 0.05 | -0.0144 | -0.0301,<br>0.0013 | 4.41 |
| Parity |  |  |  |  |  |  |
| 1 Child | Ref. |  |  | Ref. |  |  |
| 2 Children | -0.0008 | -0.0022,<br>0.0006 | 0.24 | 0.0094 | -0.0090,<br>0.0278 | -2.87 |
| 3+ Children | 0.0008 | -0.0015,<br>0.0030 | -0.23 | 0.0104 | -0.0059,<br>0.0266 | -3.17 |
| Child Died Previously |  |  |  |  |  |  |
| No | Ref. |  |  | Ref. |  |  |
| Yes | -0.0006 | -0.0016,<br>0.0004 | 0.17 | 0.0005 | -0.0027,<br>0.0036 | -0.14 |
| Perceived Baby Size |  |  |  |  |  |  |
| Large | Ref. |  |  | Ref. |  |  |
| Average | 0.0022** | 0.0006,<br>0.0039 | -0.69 | -0.0166 | -0.0424,<br>0.0092 | 5.07 |
| Small | -0.0004 | -0.0012, 0.0004 | 0.12 | -0.0032 | -0.0113, 0.0048 | 0.99 |
| Pregnancy Intent |  |  |  |  |  |  |
| Wanted Then | Ref. |  |  | Ref. |  |  |
| Wanted Later | 0.0001 | -0.0007, 0.0009 | -0.03 | -0.0001 | -0.0048, 0.0046 | 0.02 |
| Wanted No More | -0.0004 | -0.0008, 0.0001 | 0.11 | -0.0015 | -0.0046, 0.0016 | 0.46 |
| ANC Visits |  |  |  |  |  |  |
| No ANC | Ref. |  |  | Ref. |  |  |
| No ANC (Rescued) | - 0.1603**<br>* | -0.1813, - 0.1393 | 49.0<br>4 | - 0.0418**<br>* | -0.0478, - 0.0358 | 12.7<br>8 |
| 1-3 Visits | 0.0008 | -0.0034, 0.0049 | -0.23 | 0.0168 | -0.0178, 0.0515 | -5.15 |
| 4-7 Visits | 0.0004 | -0.0114, 0.0122 | -0.11 | 0.0013 | -0.0286, 0.0312 | -0.4 |
| 8+ Visits | 0 | (Omitted) | - | -0.0016 | -0.0047, 0.0015 | 0.49 |
| Mobile Phone Ownership |  |  |  |  |  |  |
| No | Ref. |  |  | Ref. |  |  |
| Yes | -0.0008 | -0.0024, 0.0008 | 0.25 | 0.0242 | -0.0242, 0.0725 | -7.39 |
| Geographical Region |  |  |  |  |  |  |
| Barisal | Ref. |  |  | Ref. |  |  |
| Chittagong | 0.0008 | -0.0004, 0.0020 | -0.25 | 0.0185** | 0.0062, 0.0309 | -5.67 |
| Dhaka | 0.0004 | -0.0005, 0.0014 | -0.13 | -0.0017 | -0.0132, 0.0098 | 0.52 |
| Khulna | -0.0001 | -0.0005, 0.0002 | 0.04 | -0.0051* | -0.0101, - 0.0002 | 1.57 |
| Mymensingh | 0 | (Omitted) | - | 0 | (Omitted) | - |
| Rajshahi | 0.0001 | -0.0003, 0.0004 | -0.02 | -0.0038 | -0.0096, 0.0020 | 1.16 |
| Rangpur | 0 | -0.0004, 0.0004 | -0.01 | -0.0055* | -0.0105, - 0.0006 | 1.7 |
| Sylhet | -0.0001 | -0.0005, 0.0002 | 0.04 | 0.0027 | -0.0008, 0.0063 | -0.84 |
| Constants |  |  |  | 0.0952 | -0.0293, 0.2197 | - 29.1<br>2 |
| Overall | - 0.2869**<br>* | -0.3096, - 0.2641 | 87.7<br>7 | - 0.0400** | -0.0638, - 0.0162 | 12.2<br>3 |
| Residual | - 0.3269**<br>* | -0.3485, - 0.3052 | 100 |  |  |  |

*Internet Access was excluded from the multivariate decomposition matrix due to near-zero*
*baseline prevalence in 2012 (1.53%). 'No ANC Provider' and 'No ANC Visits' (as well as 'First*
*Birth' and 'Parity: 1 Child') represent the same underlying unmedicalized and primiparous sub-cohorts across their respective variable classifications.*
*Coeff., Decomposition coefficients; CI, Confidence Interval; Pct., Percentage contribution of each variable category to the overall increase in Caesarean sections from 2012 to 2025; Ref., Reference. \*\*\* $p < 0.001$ , \*\* $p < 0.01$ , \* $p < 0.05$*

Among the independent variables contributing to the increase, the migration of mothers into private clinics and NGOs explained the vast majority of the compositional shift. The increased utilization of private facilities independently accounted for 43.39% of the total national surge (Coefficient: -0.1418; 95% CI: -0.1641, -0.1195; *p* < 0.001). A symmetrical reduction in home deliveries contributed an additional 42.56% to the compositional increase.

The substantial contraction of the unmedicalized demographic, evidenced by the sharp reduction in pregnancies receiving zero ANC visits, independently drove 49.04% of the structural increase (Coefficient: -0.1603; 95% CI: -0.1813, -0.1393; p<0.001). Biological indications, such as fetal macrosomia (larger than average perceived baby size), contributed less than 1% to the structural surge (Coefficient: 0.0022; *p* < 0.01), definitively ruling out changing maternal biology as a primary driver.

Sequential decompositions were executed to track the trajectory across the interim survey waves (S1 Appendix: Table S1A and S1B). During Phase 1 (2012–2019), the absorption of the unmedicalized “No ANC” cohort was the dominant driver, accounting for 58.82% of the compositional shift, alongside private facility utilization (45.73%). However, during Phase 2 (2019–2025), this unmedicalized demographic contracted to a marginal baseline due to the expansion of formal maternal care. Despite achieving this near-universal medicalization, the Caesarean rate continued to surge. During this latter phase, the private sector operated as the primary structural driver of the epidemic, independently accounting for 46.50% of the sustained surgical increase (Coefficient: -0.0747; 95% CI: -0.0849, -0.0645; *p* < 0.001).

## Discussion

This study examined the trajectory of Caesarean section deliveries in Bangladesh across 2012, 2019, and 2025, and decomposed the determinants contributing to the 32.47 percentage-point surge over this 12-year period. Although global health frameworks, such as those established by the World Health Organization (WHO), advocate for an optimal population-level Caesarean section rate of 10% to 15% to minimize maternal and neonatal mortality [1,6], the prevalence in Bangladesh reached an unprecedented 51.95% by 2025.

The multivariate decomposition analysis demonstrated that the macro-level increase was predominantly structural. At the aggregate level, 87.77% of the increase in the surgical delivery rate was attributable to compositional shifts (endowments) in the obstetric population, whereas only 12.23% was attributable to behavioral changes or shifting coefficients. Adjusting for the roles of behavioral responses, there is strong econometric evidence that the increase was driven primarily by the shift of deliveries into the private healthcare sector and the integration of previously unmedicalized populations into formal clinical pathways. Conversely, changing biological indications contributed negligibly to the trend, confirming that the rising prevalence is largely non-clinical and systemic [2,4].

The macro-level decomposition is contextualized by substantial demographic transitions within the Bangladeshi obstetric population over the 12-year study period. Descriptive analyses indicate rapid modernization, evidenced by the proportion of mothers completing secondary education increasing from 38.16% in 2012 to 54.19% in 2025 (and college or higher education rising from 14.60% to 24.88%), alongside expansions in internet access and maternal healthcare utilization. This socioeconomic transition aligns with the “obstetric transition” framework, wherein lower-middle-income countries experience large-scale shifts from home-based to facility-based deliveries [7]. However, this transition was accompanied by a pronounced homogenization of surgical rates. While Caesarean sections in LMICs have historically been concentrated among urban and wealthier populations [29], the current findings reveal a 34.83 percentage-point increase among rural women and a sixfold increase (from 5.53% to 34.37%) among the poorest wealth quintile. As maternal healthcare access expanded equitably across these demographics, the corresponding increase in surgical deliveries suggests a pattern of systemic over-medicalization that affects all strata of society, regardless of socioeconomic status or baseline clinical risk [5].

Among the independent variables contributing to the compositional increase, the utilization of private clinics and non-governmental organizations (NGOs) explained the largest proportion of the variance, accounting for 43.39% of the overall national increase. By 2025, the Caesarean section rate within private facilities had reached 85.12%. This disproportionate concentration of surgical deliveries within private facilities is further illuminated by diverging institutional trajectories. While the Caesarean delivery rate in public hospitals experienced a steady decline from 38.66% in 2012 to 33.51% in 2025, the rate within private clinics and NGOs maintained an upward trajectory, rising from 78.73% to 85.12%. This institutional divergence is a hallmark of dual-practice healthcare systems in South Asia [11]. Public facilities, often operating under high patient volumes and stringent resource constraints, tend to reserve surgical interventions for strict obstetric emergencies [30]. Conversely, the private sector absorbs the excess demand from the public system but operates within a commercialized framework where financial and operational incentives heavily favor surgical deliveries [31]. This finding aligns with systematic reviews and multi-country analyses demonstrating that private, for-profit facilities consistently exhibit the highest surgical delivery rates in low-and middle-income countries (LMICs) [2,21]. In Bangladesh, the private healthcare sector operates within a commercialized, fee-for-service environment where regulatory oversight is historically fragmented [8]. Within this market structure, financial incentives favor surgical deliveries due to higher out-of-pocket billing, predictable scheduling for obstetricians, and optimized hospital bed turnover rates [5,32]. The decomposition model indicates that if the proportion of women utilizing private facilities in 2025 had remained at the 2012 baseline level, the national Caesarean trajectory would have been substantially attenuated. This suggests that without the mandatory implementation of clinical auditing frameworks, such as the Robson classification system, private providers may continue to prioritize financial and operational factors over evidence-based maternal care [1,33].

Notably, the successful medicalization of the maternal population acted as a paradoxical secondary catalyst for the surgical surge. While the integration of vulnerable women into formal antenatal care pathways is universally recognized as a paramount public health achievement for reducing maternal mortality [18,34], the findings of this study reveal a profound medicalization paradox. As national health programs successfully encouraged the rural poor to seek formal clinical care, the healthcare architecture they entered systematically routed them toward surgical intervention. Studies across South Asia have similarly observed that increased frequency of ANC visits often exposes mothers to greater provider influence, leading to supplier-induced demand for Caesarean sections [8,22].

This supplier-induced demand is illustrated by the socioeconomic and geographic homogenization of the surgical trends. Historically, Caesarean sections in LMICs were concentrated almost exclusively among urban, wealthy, and highly educated demographics [2,14]. However, the current findings demonstrate that this surgical proliferation has rapidly expanded into previously low-intervention demographics. Between 2012 and 2025, the Caesarean section rate among the poorest wealth quintile multiplied more than sixfold (from 5.53% to 34.37%), and the surgical rate among rural women surged by 34.83 percentage points. As marginalized populations gained access to institutional delivery, severe information asymmetry between physicians and patients likely allowed providers to dictate unnecessary surgical procedures to women who lacked the health literacy or systemic empowerment to demand natural birth options [12,35].

The sequential Oaxaca-Blinder decompositions reveal that this structural escalation occurred in two distinct epidemiological phases. During Phase 1 (2012–2019), the absorption of the unmedicalized “No ANC” cohort was the dominant driver, accounting for 58.82% of the compositional shift. This represents the initial impact of integrating rural populations into the formal healthcare system. However, during Phase 2 (2019–2025), the “No ANC” demographic had been largely depleted due to near-universal medicalization. Despite this stabilization in ANC integration, the national Caesarean rate continued to rise. In this latter phase, the private sector emerged as the principal structural determinant, independently driving 46.50% of the sustained surgical increase. This phase-by-phase analysis indicates that while the initial escalation was driven by expanding access to care, the contemporary trajectory is sustained primarily by the commercial mechanisms of the private sector [36].

Furthermore, this analysis definitively rules out changing maternal biology as a primary driver of the epidemic. The decomposition results indicated that differences in perceived baby size (a proxy for fetal macrosomia) contributed less than 1% to the structural surge, while shifts in maternal age and parity were similarly negligible. This mathematically isolates the healthcare system’s structural environment, rather than the epidemiological transition of the mothers, as the root cause of the surgical escalation [4,9].

The primary strength of this study lies in the utilization of three waves of standardized, nationally representative MICS data, enabling the construction of a massive, pooled complete-case analytical cohort (N=16,179). By employing a rigorous multivariate Linear Probability Model (LPM) Oaxaca-Blinder decomposition, this analysis bypassed the path-dependence and multiplicative scaling limitations inherent to traditional logistic decompositions, resulting in exact additive percentage-point estimates for structural policy analysis [13,26]. Moreover, the methodology explicitly recoded survey skip-logic to rescue the “No ANC” cohort from listwise deletion, ensuring that the poorest and most rural unmedicalized populations were mathematically retained in the baseline architecture.

Despite these methodological strengths, several limitations must be acknowledged. First, the cross-sectional design of the MICS surveys precludes the establishment of strict temporal causality between the covariates and the surgical outcome. Second, the reliance on retrospective self-reporting for variables such as perceived baby size and delivery complications is susceptible to recall bias, although restricting the sample to births within the two years preceding the surveys mitigated this risk [19]. Most importantly, the dataset lacks clinical variables detailing the specific medical indications for surgery (e.g., fetal distress, prolonged labor, pre-eclampsia) and the direct out-of-pocket expenditures incurred by the households. Consequently, while the econometric model proves that the surge is structurally driven by private facility utilization and ANC integration, it cannot capture the localized physician-patient interactions, the degree of clinical coercion, or the specific financial devastation experienced by marginalized families [37].

## Conclusion

Over a 12-year period, Bangladesh experienced an unprecedented escalation in Caesarean section prevalence, reaching 51.95% by 2025. The present multivariate decomposition analysis demonstrated that this surge was overwhelmingly structural, driven primarily by the disproportionate concentration of deliveries within the private healthcare sector and the near-universal integration of previously unmedicalized populations into formal antenatal care pathways. Conversely, shifting biological indications contributed negligibly, confirming a profound pattern of systemic over-medicalization independent of baseline clinical risk. Reversing this medically unjustified trajectory necessitates immediate regulatory intervention. Public health policies must mandate clinical auditing frameworks, such as the Robson classification, to objectively monitor surgical indications and decouple commercial financial incentives from obstetric decision-making in private facilities [32,33]. Ultimately, while sustaining equitable maternal healthcare access remains a public health priority, future health system strategies must explicitly integrate protocols that protect the physiological integrity of childbirth against supplier-induced surgical demand [2].

## Acknowledgments

The author gratefully acknowledges the United Nations Children’s Fund (UNICEF) and the Bangladesh Bureau of Statistics (BBS) for the design, implementation, and provision of the Multiple Indicator Cluster Surveys (MICS) data utilized in this study.

## Author Contributions

**Conceptualization**: Md Akhtaruzzaman Limon.

**Data curation**: Md Akhtaruzzaman Limon.

**Formal analysis**: Md Akhtaruzzaman Limon.

**Methodology**: Md Akhtaruzzaman Limon.

**Writing - original draft**: Md Akhtaruzzaman Limon.

**Writing - review & editing**: Md Akhtaruzzaman Limon.

## Data Availability Statement

The datasets generated and analyzed during the current study are available in the UNICEF Global Multiple Indicator Cluster Surveys (MICS) repository. Access to the raw data is freely available to researchers upon registration and approval via the official MICS portal (https://mics.unicef.org/surveys).

## Supporting information

S1 Appendix. Sequential Oaxaca-Blinder decompositions for Phase 1 (2012–2019) and Phase 2 (2019–2025). This file contains Table S1A and Table S1B detailing the absolute percentage-point contributions of compositional and behavioral shifts across the interim survey waves.

